# EVALUATION OF ENVIRONMENTAL AND SOCIOECONOMIC FACTORS CONTRIBUTING TO FRAGILITY FRACTURES IN INDIANS

**DOI:** 10.1101/2020.06.23.20135178

**Authors:** Vaibhav Singh, Ananda Kisor Pal, Dibyendu Biswas, Alakendu Ghosh, Brijesh P Singh

## Abstract

**Objective:** Osteoporosis causes fragility fractures that also occur in patients with bone mineral density (BMD) in the normal or osteopenic range, suggesting role of risk factors that are unrelated or partially related to BMD. The study aims at highlighting the link between 3 conditions, that are environment and occupation related risk factors and that are widely prevalent in India, and development of fragility fractures.

**Methods:** A Case Control study was done by recruiting 110 Cases with history of recent fragility fractures and 84 Controls with no history of recent fractures. 3 study parameters, village dwelling, conventional farming, and poverty, were chosen the presence or absence of which were documented in participants. This was followed by an ODDS ratio analysis.

**Results:** The Odds of village dwellers, conventional farmers, and socioeconomically poor individuals to develop fragility fractures were both significant and large.

**Conclusion:** Urbanization is a risk in the development of fragility fractures. However, this study points that village dwelling in India is associated with the development of fragility fractures. Similarly, Odds of farmers exposed to pesticides and agrochemicals to develop fragility fractures is large and significant. Pesticides and agrochemicals act as endocrine disruptors and bone health is closely linked to endocrine system. Fragility fractures among farmers may be due to endocrine disrupting properties of pesticides and agrochemicals. Socioeconomic deprivation is a known risk in the development of osteoporosis. This study too highlights that the odds of individuals living in poverty to develop fragility fractures is significant and large.

## INTRODUCTION

Osteoporosis can be defined as a metabolic bone disease distinguished by such characteristics as reduced bone mass, microarchitectural deterioration of bone tissue, and an increased risk of fragility fractures.[1] The diagnosis of osteoporosis is made in individuals with bone mineral density (BMD) of 2.5 or more standard deviations (SD) below the mean for young adult reference population (T score ≤ −2.5).[2] Individuals with lesser reductions in BMD (T score between −1 and −2.5) are considered osteopenic.[2] T scores between −1 and 2.5 are considered normal.[2] Osteoporosis leads to fragility fractures that occur spontaneously or due to low intensity trauma such as fall from standing height or fall from bed. Fragility fractures occur commonly at hip, spine, wrist, and proximal humerus. A large number of fragility fractures occurs in individuals whose BMD is in the osteopenic range.[2] BMD is just 1 component of fracture risk, and there are other risk factors and abnormalities in skeleton that contribute to fragility.[3] There are numerous risk factors that eventually lead to fragility fractures.[4] Some risks act independent of BMD whereas certain other risks are thought to influence BMD. One of the observations in our routine clinical practice was that a number of fragility fractures occurred in patients with osteopenia or normal BMD. This made one argue that some risks were probably unaccounted for up till now that might act independent of BMD. Such fragility fractures, occurring in the backdrop of normal BMD or osteopenia, hint at risk factors that act independently of BMD and that are different from clinical risk factors included in fracture risk assessment. To peep into the underlying risks associated with development of fragility fractures in Indian population, we identified 3 conditions that were widely prevalent in India, that were related to surrounding environment and lifestyle, and that were not captured individually in fracture risk assessment normally employed in routine clinical practice. These 3 conditions were village dwelling, conventional farming, and poverty. India predominantly lives in villages. Urbanization is a risk factor for fragility fractures and osteoporosis due to changing lifestyle and diminished physical activity.[5,6] India is an agrarian country with majority of Indians involved in farming and other related activities. Widespread use of pesticides and other agrochemicals for farming is prevalent in India.[7] Subsequent exposure of Indian farm families to pesticides and other agrochemicals appears logical. The effect of exposure to pesticides and agrochemicals on bone health is still largely unknown and ought to be deciphered. Many pesticides available commercially disrupt the endocrine system of the human body,[7] and optimum bone health is closely linked to proper functioning of the endocrine system.[2] Osteoporosis and fragility fractures are common in socially deprived individuals.[2] Through this Case Control Study conducted at IPGME&R and SSKM hospital, Kolkata, we intended to answer whether village dwelling conferred protection from fragility fractures and whether exposure to pesticides and agrochemicals, while practicing farming as an occupation, and low socioeconomic status were associated with the development of fragility fractures.

## MATERIALS AND METHODS

IPGME&R and SSKM hospital, Kolkata treats patients who reside in many states of eastern India. Moreover, Kolkata is a cosmopolitan with residents who are native of different regions of the country. Data was collected from participants from June 2017 to April 2019. Cases and Controls were selected on the basis of preset Inclusion and Exclusion criteria.

### Inclusion Criteria for Cases Group

1. Recent history of fragility fracture-fragility fractures were defined as those occurring spontaneously or those occurring due to fall from standing position or fall from bed.
2. Age: 40-90 years
3. Appropriate history of Clinical Risk Factors of Osteoporosis be made available along with occupational history, history of exposure to pesticides and agrochemicals if occupation is related to farming, information about whether residence is in rural area or urban area, and appropriate information about cumulative family income and total number of family members
4. Report of investigations ordered at first presentation be made available within 30 days of date of fracture
5. Site of fracture: Proximal humerus, wrist, spine, and hip
6. No history of bisphosphonate, teriparatide, or other anti-osteoporotic pharmacotherapy

### Inclusion Criteria for Controls Group

1. No history of recent fracture
2. Age 40-90 years
3. Appropriate history of Clinical Risk Factors of Osteoporosis be made available along with occupational history, history of exposure to pesticides and agrochemicals if occupation is related to farming, information about whether residence is in rural area or urban area, and appropriate information about cumulative family income and total number of family members
4. Report of investigations ordered at first presentation be made available within 30 days of date of fracture
5. No history of bisphosphonate, teriparatide, or other anti-osteoporotic pharmacotherapy

### Exclusion Criteria

1. Road traffic accidents and high intensity trauma
2. Age<40 or Age>90
3. Low intensity trauma fractures at sites other than that at hip, spine, proximal humerus, or wrist
4. Pathological fractures
5. Investigations ordered were either not done or done more than 30 days after the date of fracture in case of Case group

After the study proposal was approved by the Institutional Ethics Committee, a total of 110 Cases and 84 Controls were enrolled randomly for the purpose of this Institution Based Case Control Study. The participants were made aware of the nature of this study along with requirements for enrolment in the study. A written informed consent was received by participants before formal enrolment in the study. A total of 7 patients of proximal humerus fractures, 24 patients of hip fractures, 44 patients of vertebral compression fractures, and 35 patients of wrist fractures were included in the Case Group after these patients met the inclusion criteria. Data under the following headings was collected from all participants:

1. Name
2. Site of Injury in Case group
3. Age
4. Sex
5. Occupation
6. If Occupation is Farming then General Details of Crops Grown
7. History of Exposure to Pesticides and Agrochemicals in case of farmer
8. Cumulative Family Income along with Total Number of Family Members
9. Bilateral Femoral Neck BMD through DEXA Scan
10. Serum 25(OH)D3
11. Radiography
12. Serum Calcium
13. Serum Phosphate
14. Serum Alkaline Phosphatase

Radiography of the fracture site was done in Cases to confirm the diagnosis. Some Controls were subjected to radiography to rule out fractures-in case the control complained of chronic back pain or chronic pain at some other site. Radiography was not done in CONTROLS who were asymptomatic with apparently healthy bones to avoid radiation exposure in them. Serum 25(OH)D3, Serum Calcium, Serum Phosphate, and Serum Alkaline Phosphatase were done mainly to rule out common causes of pathological fractures other than osteoporosis such as bone metastasis, osteomalacia, renal osteodystrophy, primary and secondary hyperparathyroidism.

The disease-Fragility Fracture-was present in Cases and absent in Controls. Presence or absence of 3 potential conditions were identified in every participant; these 3 conditions were conventional farming, village (rural area) dwelling, and low socioeconomic status. An Odds ratio analysis was done, through binary logistic regression function of IBM SPSS Statistics 21 software, at 95% confidence interval.

Odds of participants dwelling in villages of developing fragility fractures was calculated with respect to those dwelling in urban areas. A substantial proportion of participants of the study were conventional farmers and were exposed to pesticides and agrochemicals. All the farmers who participated in the study were conventional farmers. Odds of participants associated with conventional farming of developing fragility fractures was calculated with respect to those not associated with it. Similarly, participants were again divided into three groups based on incomes per member of family per month, which was calculated by dividing total family income by total number of family members. The World Bank defines extreme poverty as dollar earnings of less than $ 1.90 per day per person.[8] The sum of $1.90 was roughly equivalent to Rs 134 based on exchange rates on November 29, 2018. Using this as a rough guide, we arbitrarily divided participants in to 3 groups-those sustaining on ≤ Rs 3000/month per person (extreme poverty group), those sustaining on ≥ Rs 3001/month per person and ≤ Rs 6000/month per person (moderate poverty group), and those sustaining on ≥ Rs 6001/month per person (not associated with poverty). The Odds of the extreme poverty group and the moderate poverty group of developing fragility fractures were calculated with respect to those not associated with poverty.

## RESULTS

### Age

Age distribution of Cases and Controls is depicted in Figure 1.

**FIGURE 1:**
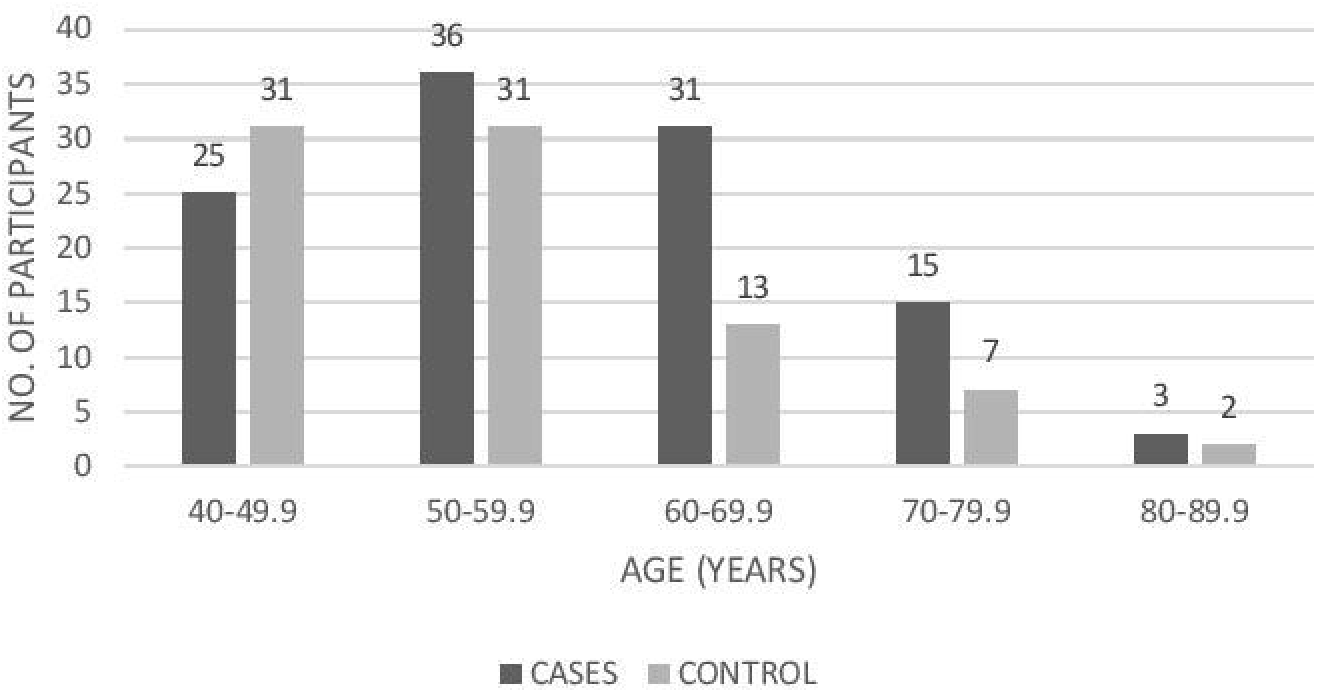
AGE DISTRIBUTION OF CASES AND CONTROLS

### Sex

Sex distribution of Cases and Controls is described in Figure 2.

**FIGURE 2:**
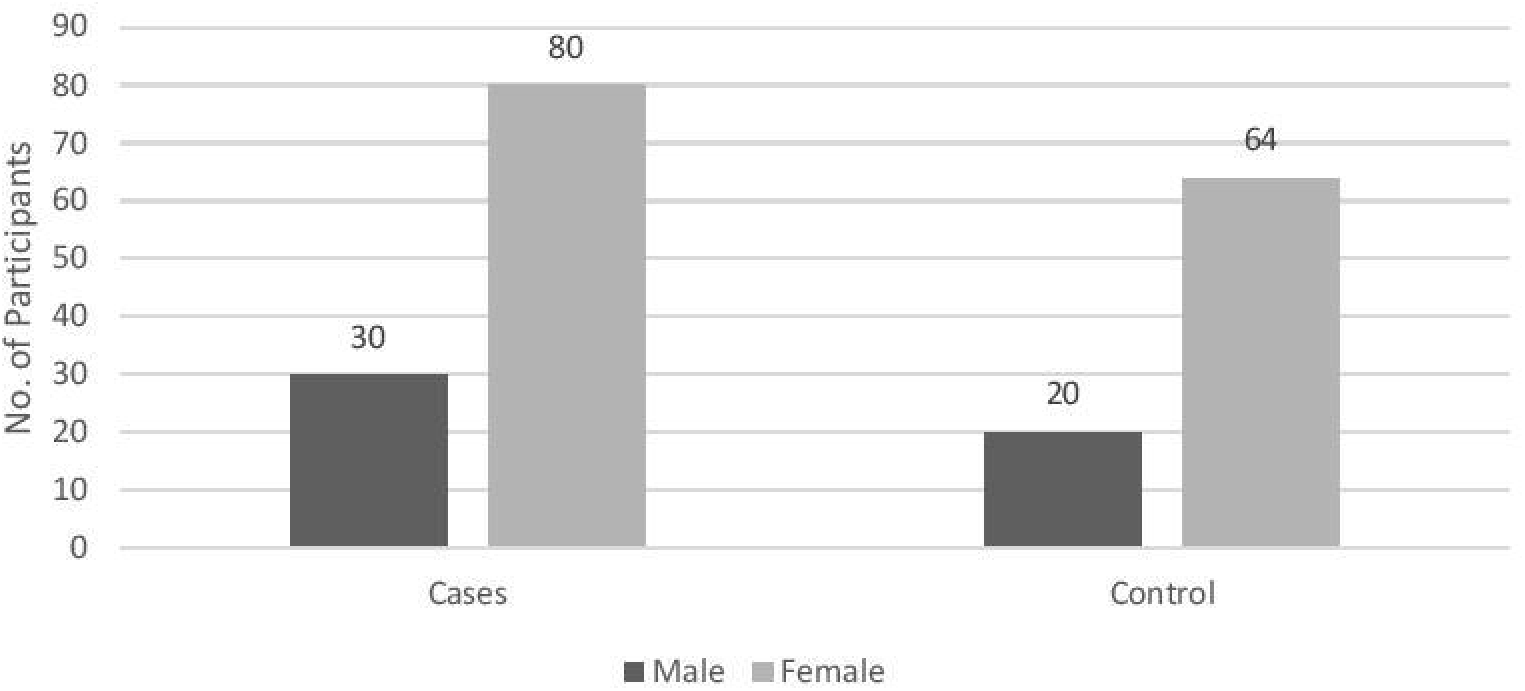
SEX DISTRIBUTION OF CASES AND CONTROLS

### Bone Mineral Density

Bone mineral density (BMD) of Cases and Controls were documented. Using this BMD, a T score was calculated using NHANES III female reference data. Cases and Controls were stratified into 3 groups-Osteoporotic, Osteopenic, and Normal-as described in Figure 3.

**FIGURE 3:**
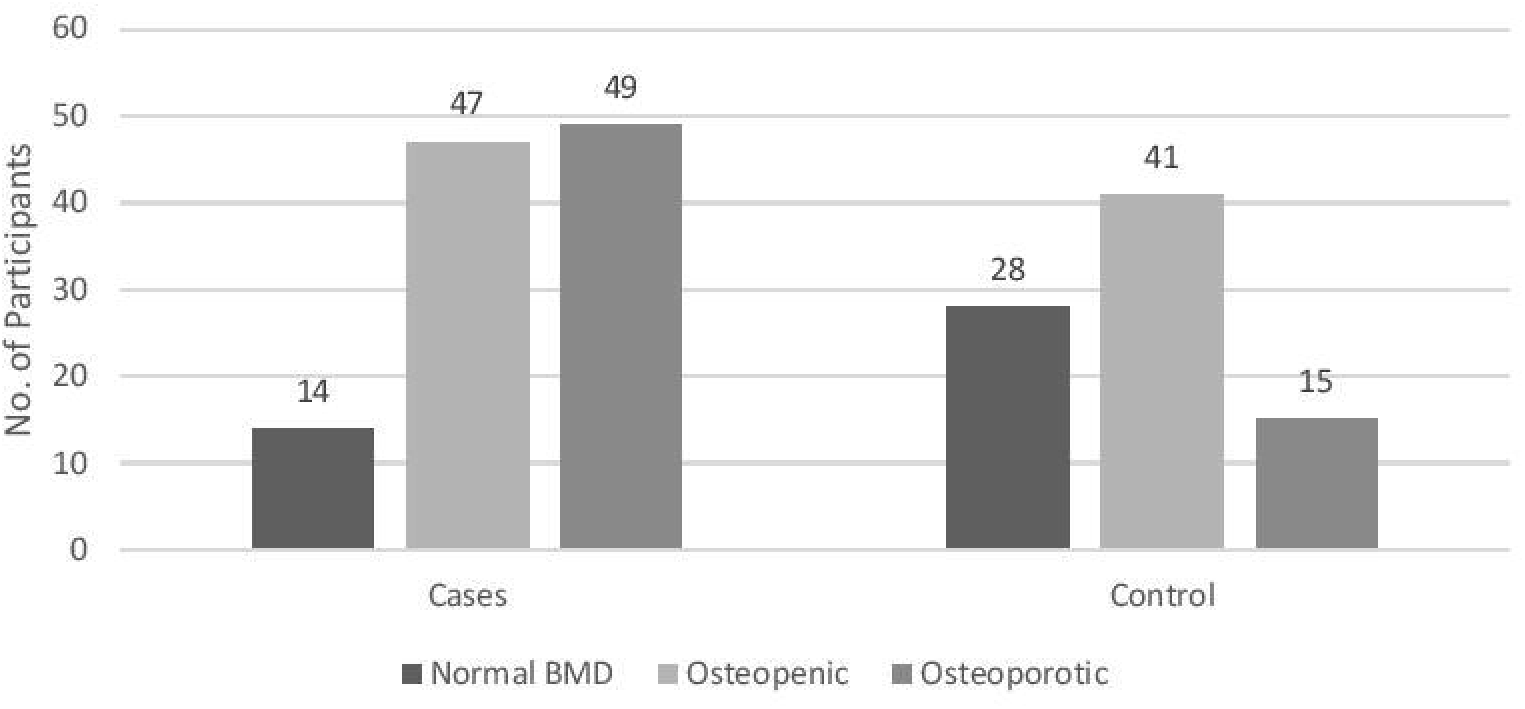
BMD DISTRIBUTION OF CASES AND CONTROLS

### Village Dwelling

59.09% of Cases and 39.29% of Controls resided in villages. The Odds of village dwellers to develop fragility fractures was 2.232 times compared to urban dwellers, a significant risk as shown in Table 1.

**Table 1:** Odds Ratio of Risk Factors

| Table 1: Odds Ratio of Risk Factors |  |  |  |  |
| --- | --- | --- | --- | --- |
|  | ODDS RATIO | P VALUE | 95% CONFIDENCE INTERVAL |  |
|  |  |  | LOWER LIMIT | UPPER LIMIT |
| Village (Rural)<br>Dwelling | 2.232 | 0.007 | 1.250 | 3.986 |
| Conventional Farming | 2.353 | 0.010 | 1.232 | 4.493 |
| Extreme poverty | 3.597 | 0.001 | 1.667 | 7.758 |
| Moderate poverty | 4.062 | 0.012 | 1.356 | 12.172 |

### Conventional Farming

39.09% of Cases and 21.43% of Controls practiced conventional farming as a means of living and were exposed to pesticides and agrochemicals more than the other participants who were non farmers. The Odds of conventional farmers to develop fragility fractures was 2.353 times compared to non-farmers-the risk was significant at 95% confidence interval as shown in Table 1.

### Poverty

Participants were divided into 3 groups based on their incomes per month per family member as shown in Figure 4. This was calculated by total family income divided by total number of family members. The ‘Extreme Poverty’ group comprised of those individuals whose income per family member was less than Rs 3000/month. The ‘Moderate Poverty’ group comprised of those individuals whose income per family member was between Rs 3000/month and Rs 6000/month. Participants whose income per family member was greater than Rs 6000/month comprised the ‘Not Poor’ group. The Odds of Extreme Poverty group and Moderate Poverty group to develop fragility fractures was calculated with respect to Not Poor group. The Odds of Extreme Poverty Group to develop fragility fractures was 3.597 and the Odds of Moderate Poverty Group to develop fragility fractures was 4.062 compared to Not Poor Group, the result being significant at 95% confidence interval (Table 1).

**FIGURE 4:**
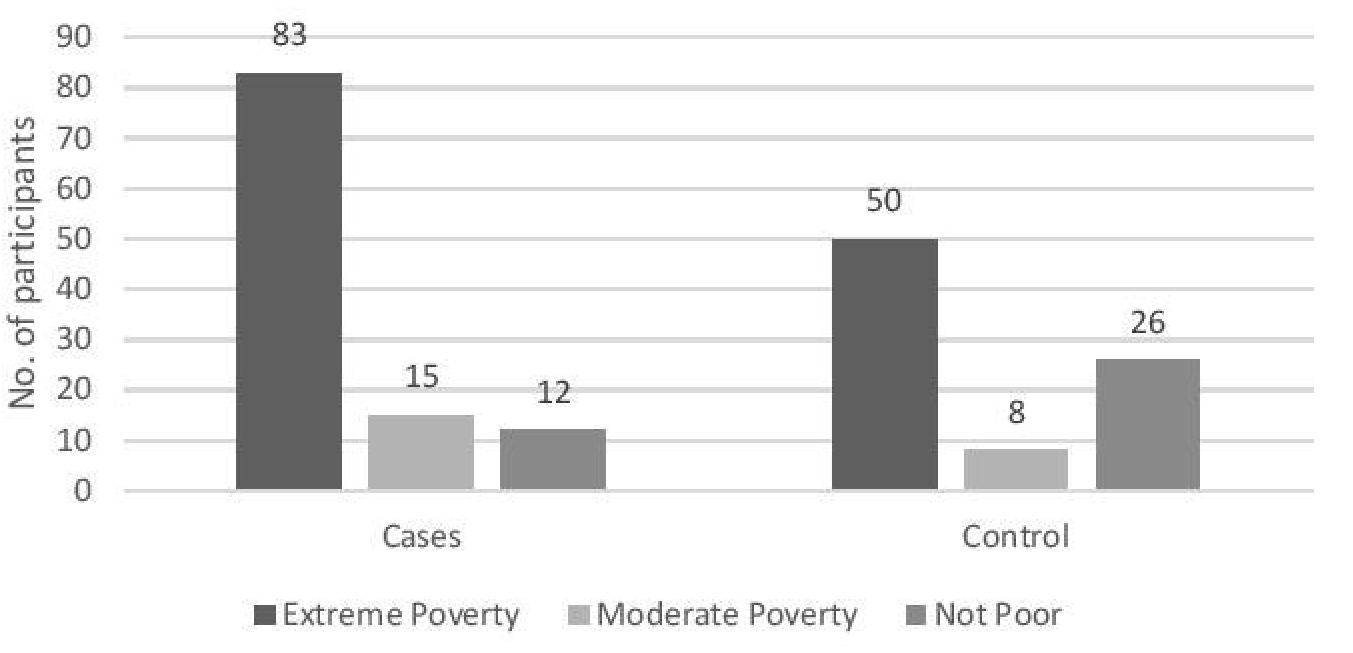
SOCIOECONOMIC DISTRIBUTION OF CASES AND CONTROLS

## DISCUSSION

Although, cases were mostly between the ages 40 years and 70 years, a significant number of fragility fractures occurred in the younger age groups, that is, individuals younger than 50 years of age. The risk of fragility fractures increases with advancing age.[9] A large number of fragility fractures occurring in relatively younger population probably points at risk factors other than advancing age at play. The majority of participants in this study, both in the Cases group and in the Controls group, was female. Participants in the Cases group who suffered a recent fragility fractures had their BMDs mostly in the osteopenic and the normal ranges although a large number of participants in the Cases group had osteoporotic BMDs. Fragility fractures occurring in the backdrop of normal BMD or osteopenic BMD pointed at risk factors that were unrelated or partially related to changes in BMD. The Geelong Osteoporosis study, a population-based study on osteoporosis in Australia, suggested a lower rate of hip fractures among rural dwellers compared to urban residents and a lower rate of fractures associated with osteoporosis among rural dwellers compared to urban residents.[10] A descriptive study found that fracture rates were higher among urban residents of central city of Rochester compared to those of rural residents of Olmsted county, Minnesota.[11] Similarly, a population-based study in Southern Sweden revealed higher relative risk of fractures among urban residents compared to their rural counterparts, especially in the elderly.[12] A study done in Hongkong suggested a substantial increase in the age specific rate of hip fractures from 1965 to 1985, although no increase in age specific rate of hip fracture was documented from 1985 to 1995.[13] A population-based study done in Singapore noted an increase in incidence of hip fracture, a site common for osteoporotic fracture, from 1960s to 1991-1998.[14] A Japanese study documented that age specific incidence rates of hip, distal radius, and proximal humerus fracture increased between the observed periods 1992-1994 and 2010-2012.[15] The Hongkong, Singapore, and Japanese studies suggested an association between increase in hip fracture rates and rapid urbanisation, which was accompanied by changes in nutrition and the level of physical activity.[5,6] According to our analysis, the Odds of village dwellers to develop fragility fractures was 2.232 times that of urban dwellers and the result was statistically significant at 95% confidence interval. Traditionally, rural and village areas are thought to have clean environments compared to towns and cities, and rural environments are considered good for human health. Residents of rural areas are considered having increased levels of physical activity that is thought to provide protection against fractures. However, our study points to the contrary. According to our study, village dwellers in India are at increased odds to develop fragility fractures compared to urban dwellers. In addition to the environment, this trend may be linked to exposure to other environmental toxins.

Rural areas in India are immensely linked to farming and related activities. Traditional methods of farming are increasingly being replaced by Indian farmers in favour of modern conventional methods of farming that incorporate the use of pesticides, fertilizers, and other agrochemicals. Conventional farming adapted by Indian farmers is not only chemical intensive but also capital intensive and energy intensive.[7] Pesticides and agrochemicals are marketed in India as “medicine for the plants”.[7] The Indian farmers and peasants usually have low awareness of the hazard potential of pesticides and agrochemicals, the dosage protocols, and the safety measures. Farm activities such as spraying of pesticides and application of fertilizers are carried out without the use personal safety gear.[7] As a result, exposure to pesticides and agrochemicals is probably rampant and undocumented in India. Pesticides and agrochemicals are known to be hazardous to human health. Non-judicious, excessive, and improper use of pesticides and agrochemicals intuitively may bring health risks to its user and to the larger community exposed to it. The next logical enquiry in this study was to decipher whether conventional farming and related activities were associated with the development of fragility fractures. All participants of our study, except one, who were associated with farming activities resided in villages. All the participants who practiced agriculture for a living were exposed to pesticides and other agrochemicals. None of them gave history of using personal protective gears while handling pesticides and agrochemicals. Many of the participants who were farmers stored these pesticides and agrochemicals in homes. They never discarded their clothing after pesticide and agrochemical application; often pesticide laden clothing was brought home to be washed by hand by other members of the family who also got exposed through it. Many of the participants who were farmers were small family farm owners with very little education or no formal education. This might have added to the exposure risk. The duration of pesticide and agrochemical exposure was long too often continuing for years or decades. The Odds of conventional farmers to develop fragility fractures was 2.353 times that of nonfarmers and the result was statistically significant at 95% confidence interval. Conventional framing is the term broadly used to describe the agriculture practices that are a product of Green Revolution. These supposedly modern agriculture practices make rampant use of chemical fertilizers and pesticides including herbicides, insecticides, rodenticides, fungicides, molluscicides, etc. Farmers and bystanders are exposed to pesticides in a number of situations such as mixing, application, sale, transportation, storage, maintenance of equipment, spillage, re-entering farms, disposal, etc.[16] India is an agrarian country with a large chunk of its population involved with farming and related activities. There is a substantial body of evidence that suggests that pesticides in the ecosystem disrupt the endocrine system. The effects of pesticides on the endocrine system mimic those of endocrine disruptors. Endocrine disruptors have been broadly defined as exogenous agents that interfere with production, release, transport, metabolism, binding, action, or elimination of natural hormones in the body responsible for maintenance of homeostasis and the regulation of developmental process.[17] At a cellular level, endocrine disruption refers to a mechanism of toxicity that interferes with the ability of the cells to communicate hormonally and results in a wide variety of adverse health effects including birth defects, reproductive, developmental, metabolic, immune, and neurobehavioral disorders as well as hormone dependent cancers.[18] A study showed that trifluralin, triadimefon, parathion, malathion, methomyl, carbaryl, aldicarb, dicofol, ziram, maneb, mancozeb, vinclozolin, iprodione, and benomyl were some chemicals used in agriculture that were known to cause endocrine disruption and that were associated with neural tube defects in new born of mothers residing within 1000 m of farms using these chemicals.[19] Alachlor, metribuzin, and parathion are possible endocrine disruptors affecting estrogen, androgen, thyroid hormones, progesterone, follicle stimulating hormone, and luteinizing hormone metabolism, and there are other estrous cycle disruptors used in agriculture such as carbaryl, carbofuran, cyanazine, parathion, and petroleum oil.[20] A number of the above mentioned pesticides, especially parathion and malathion, are a commercial success in the Indian market. Due to a substantial body of evidence that has surfaced pointing at the endocrine disrupting properties of several agricultural pesticides, Pesticide Action Network, UK has listed 101 pesticides as proven or possible endocrine disruptors in 2009.[21] There are several diseases of the endocrine system that are related to development of osteoporosis and fragility fractures such as early menopause, thyrotoxicosis, primary hyperparathyroidism, cushing syndrome, and hypogonadism.[2] Use of a few hormonal drugs are also implicated in the development of osteoporosis and fragility fractures such as use of corticosteroids, thyroxine, and gonadotrophin releasing hormone agonist.[2] Several hormonal agents such as parathyroid hormone, calcitonin, calcitriol, testosterone, hormone replacement therapy (HRT) are used in the treatment of osteoporosis.[2] Thus, it is possible, theoretically, that exposure to an exogenous substance that can potentially disrupt the endocrine system generally and androgen, estrogen, and thyroid hormone systems specifically can adversely affect the bone health leading to fragility fractures.

Social deprivation is known to have a role in the development of fragility fractures.[2] According to a retrospective study, an increment of US$ 10000 in GDP per capita was associated with 1.3% increase in hip fracture probability; this might be due to the fact that socioeconomic prosperity might lead to diminished levels of physical activity and increased chances of falling on hard surfaces.[22] On the contrary, another retrospective study concluded that hip fracture rates decreased with increasing income.[23] A population-based UK study also found strong associations between deprivation and fracture risk in men such as risk of hip, wrist, and vertebral fractures; the relative risk was greatest for hip fractures.[24] A retrospective population-based US study concluded that low income populations were at increased risk of hip fractures.[25] Another UK study also suggested a significant 1.3 fold increase in the incidence of hip fractures among the most deprived population compared to the least deprived.[26] A retrospective Portuguese study exhibited an increased risk of hip fractures in individuals of both sexes residing in deprived municipalities compared to those residing in more affluent municipalities.[27] An observational cross-sectional study concluded that postmenopausal women living in poverty have a lower BMD at lumbar spine and a higher prevalence of osteoporosis compared to women not living in poverty.[28] According to our analysis, the Odds of individuals in extreme poverty group (those earning less than Rs 3000 per month per person) to develop fragility fractures was 3.597 times and the odds of individuals in the moderate poverty group (those earning between Rs 3000 and Rs 6000 per month per person) to develop fragility fracture was 4.062 times compared those who earn more than Rs 6000 per month per person-the result being statistically significant at 95% confidence interval. The result is consistent with the majority of existing literature on the effects of socioeconomic status on the development of osteoporosis and fragility fractures. This study has its limitations. The sample recruited in this study has a moderate size. The investigations ordered for the purpose of this study were not done from 1 laboratory. Investigations were done from both IPGME&R and private laboratories. As a result, their was difference in the machines used for investigations and their maintenance status. Nevertheless, it is important to mention that all Dexa scans, to evaluate BMD, were done by machines that belonged to the manufacturer, GE Lunar.

## CONCLUSION

Village dwelling, conventional farming, and low socioeconomic status are associated with development of fragility fractures in India. The study also highlights the need for further research to accurately decipher the link between conventional farming, village dwelling, and socioeconomic status and bone health and the role of each of these risks in the development of osteoporosis and fragility fractures.

## Data Availability

The data that helped us reach conclusion of this study is available on www.osf.io
Identifier: DOI 10.17605/OSF.IO/HMU2W
The authors have ownership of primary patient data which is available for review by concerned authorities upon request.

https://www.doi.org/10.17605/OSF.IO/HMU2W

## ACKNOWLEDGEMENT

I feel deep gratitude for the following teachers, mentors, and supporters without whom this study would not have met the desired outcome.

1. Dr (Prof) K Chakraborty, Department of Orthopaedics, IPGME&R & SSKM Hospital, Kolkata.
2. Dr T. Dutta, Department of Orthopaedics, IPGME&R & SSKM Hospital, Kolkata.
3. Dr A. Karmakar, Department of Orthopaedics, IPGME&R & SSKM Hospital, Kolkata.
4. Dr (Prof) GK Dhali, Chairman, Research Oversight Committee, IPGME&R & SSKM Hospital, Kolkata.
5. Dr M. Mukhopadhyay, Head, Department of Biochemistry, IPGME&R & SSKM Hospital, Kolkata.
6. Dr (Prof) SK Mishra, Department of Mathematics, Institute of Science, Banaras Hindu University, Varanasi.
7. Dr (Prof) KK Singh, Department of Statistics, Institute of Science, Banaras Hindu University, Varanasi.

## DECLARATIONS

### Ethics approval

The study was based on the thesis submitted for successful completion of the degree MS (Orthopaedics) from The West Bengal University of Health Sciences, Kolkata, India. The study was also approved by the Institutional Ethics Committee of IPGME&R and SSKM hospital, Kolkata. The certificate is attached underneath.

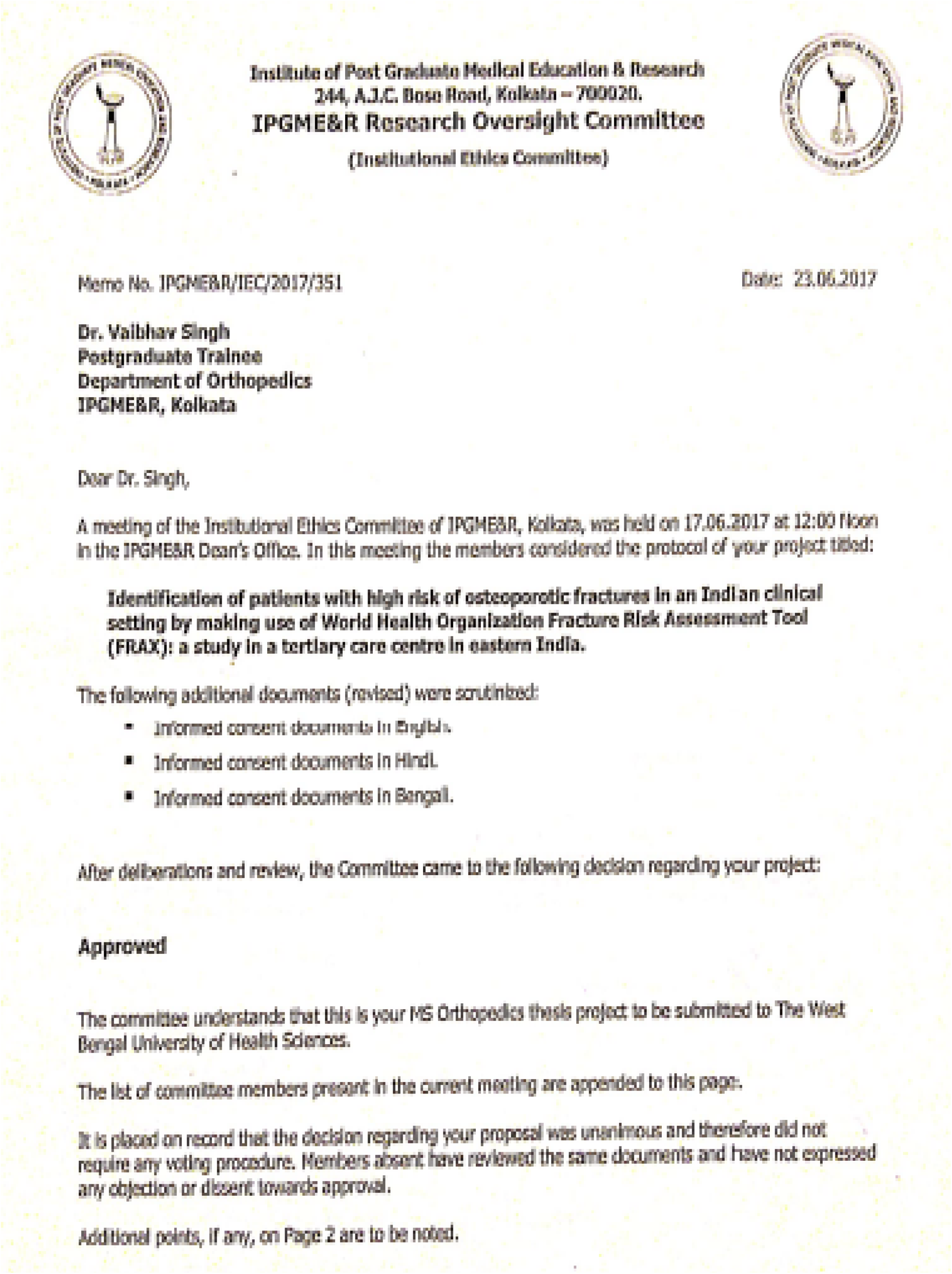

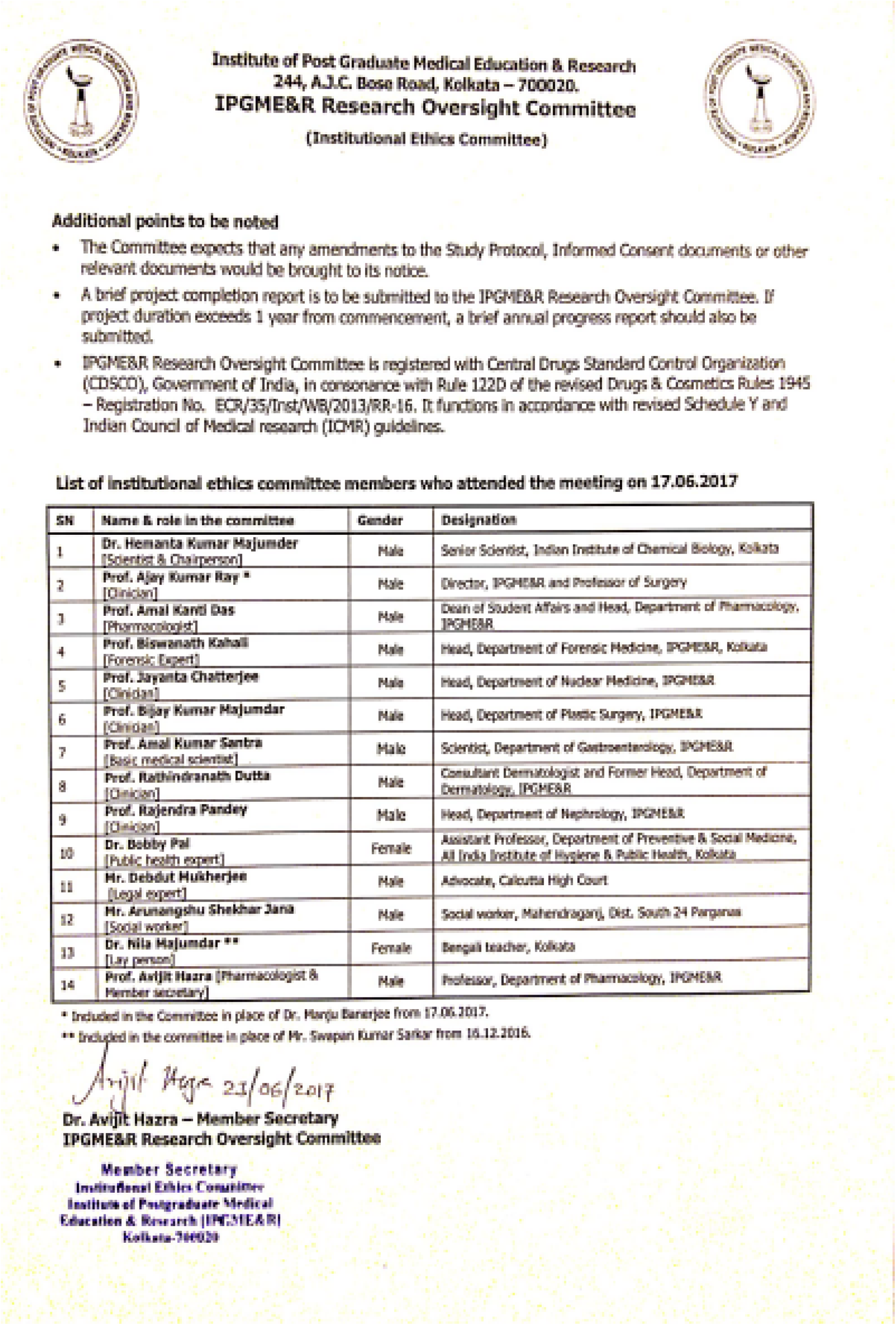

### Consent to participate

All the participants of the Case group and the Control group were explained the nature and scope of the study. They were made aware of all the requirements necessary to participate in the study. All participants signed a consent form before formal enrolment in the study.

### Consent for publication

All authors, as well as their respective Departments and Institutions, agree to publish the findings of the study.

### Availability of data and material

The data that helped us reach the outcome of this study is available on https://www.doi.org/10.17605/OSF.IO/HMU2W

The authors have ownership of the primary patient data that is available for review by concerned authorities.

